# APRI score is not predictive of post-surgical outcomes in cholangiocarcinoma patients

**DOI:** 10.1101/2023.02.12.23285817

**Authors:** Faaiq N Aslam, Tristan A Loveday, Pedro Luiz Serrano Uson Junior, Mark Truty, Rory Smoot, Tanios Bekaii-Saab, Daniel Ahn, Mohamad Bassam Sonbol, Christina Wu, Chee-Chee Stucky, Hani Babiker, Mitesh J Borad

## Abstract

**Objective:** To assess the utility of aminotransferase-platelet ratio index (APRI) score in prognosticating post-surgical outcomes in cholangiocarcinoma patients.

**Methods:** 152 cholangiocarcinoma patients at the Mayo Clinic that underwent surgical resection were retrospectively analyzed. Statistical analyses were done to determine the relationship between APRI score and demographic factors, laboratory values, pathological features, and outcomes data.

**Results:** No relationship between APRI score and demographic factors was identified. There was a correlation between APRI score and ALT, albumin, and bilirubin, but the remaining laboratory parameters showed no correlation. APRI score did not prove to be useful as a prognostic tool as it did not correlate with tumor pathology features and did not correlate with post-surgical recurrence and mortality.

**Conclusion:** We found that APRI score was not a prognostic tool for post-surgical outcomes in cholangiocarcinoma patients.

## Introduction

Cholangiocarcinoma is the second most common primary liver cancer. Tumors can arise at different sites (intrahepatic, extrahepatic, perihilar), from different cells of origin, and are genomically and histologically heterogeneous.^1, 2^ The pathogenesis of cholangiocarcinoma is felt to largely be driven by chronic inflammation, and cholestasis which subsequently results in cellular proliferation, genetic and epigenetic changes, and eventually carcinoma.^3^ Conditions that increase inflammation like hepatitis B, hepatitis C, liver cirrhosis, primary sclerosing cholangitis (PSC), non-alcoholic fatty liver disease (NAFLD), non-alcoholic steatohepatitis (NASH) and liver flukes are well known risk factors for cholangiocarcinoma.^2, 4-6^

As with many malignancies, treatment options for cholangiocarcinoma include surgical resection, chemotherapy, targeted therapies, immunotherapy, and radiation therapy, with surgery/transplant being the only curative option.^2, 4^ However, surgical resection outcomes still leave much to be desired. Five-year survival rates remain around 20-40%.^2, 4, 7-9^ Liver transplant is only an option with selected cases of perihilar disease or early intrahepatic disease and can achieve five-year survival rates of around 65-70%.^2, 4^ Factors that are related to 5-year mortality/survival include lymph node status, margins status, histological grade, vascular invasion, and tumor size.^2, 5, 7, 8^ However, these prognostic indicators are only known after surgical resection. Unfortunately, although the utility of CEA and CA19-9 levels have been studied, studies have yielded mixed results and there are no definitive pre-operative prognostic implications.^5, 10^ A reliable pre-surgical prognostic indicator for post-resection outcomes could be extremely valuable in surgical decision making and post-operative prognostic guidance for these patients.

Given that the underlying disease processes of cholangiocarcinoma are related to chronic inflammation of the liver and biliary tree, indicators of liver inflammation and fibrosis could be useful in assessing cholangiocarcinoma outcomes.^3^ The APRI score, calculated as the ratio of serum AST to platelet count, has shown to be a useful marker for liver fibrosis.^11^ It was first implemented as a simple, non-invasive indicator of liver fibrosis and cirrhosis in chronic hepatitis C patients.^12^ However, its utility has subsequently expanded beyond hepatitis C patients.^11^ For example, as a result of its utility as a useful marker for fibrosis, APRI scores have shown to correlate with post-surgical liver failure and mortality in patients undergoing resection for hepatocellular carcinoma (HCC) and for mixed HCC-cholangiocarcinoma tumors.^13^ Given these findings, we hypothesized that APRI score could potentially be a useful marker for measuring disease severity and prognosticating disease in cholangiocarcinoma patients. Therefore, we performed a retrospective analysis of 152 cholangiocarcinoma patients at the Mayo Clinic who underwent surgical resection to determine whether an array of patient and tumor characteristics correlated with APRI score and whether pre-operative APRI scores could be used to predict post-surgical outcomes in cholangiocarcinoma patients.

## Methods

### Data Collection

This study entailed a retrospective analysis of 152 patients at the Mayo Clinic who had cholangiocarcinoma and underwent surgical resection between 2010 and 2020. Patient demographic data, laboratory parameters prior to surgery, tumor pathology, and outcome data were obtained. Demographic data included date of age, sex, race, tumor type, and fibrosis etiology. Laboratory data included CA19-9, albumin, bilirubin, international normalized ratio (INR), alkaline phosphatase, alanine aminotransferase (ALT), aspartate aminotransferase (AST), and Platelet Count. Child Pugh and APRI scores were calculated using the laboratory parameters listed and clinical data for ascites and encephalopathy for Child Pugh score. Pathology data included tumor size, status of vascular and/or perineural invasion, tumor grade, tumor stage, tumor margin status, and margin width. Outcome data included recurrence status, time to recurrence, vital status, and survival from time of surgery. The data was subsequently analyzed to determine if there was a relationship between APRI score and the demographic, laboratory, pathology, and outcome data, respectively. Approval from the Mayo Clinic institutional review board was obtained prior to data collection for this study.

### Data Analysis

The data was analyzed using the SciPy stats package and figures were generated using the MatPlotLib package for Python.^14, 15^ To determine the relationship between quantitative laboratory, pathology, and outcome data, datapoints for each measure and the associated APRI score were plotted on a scatter plot. Then, using the SciPy stats package, the Pearson correlation coefficient, p-value (with p < 0.05 being considered as statistically significant), line of best fit, and slope were determined.

For qualitative demographic and pathology data, box plots were generated to illustrate how APRI score varied in each category. Then, depending on the number of variables, a 2-sample t-test or a one-way ANOVA test were done using the SciPy package to determine whether APRI score is significantly different among the different variables (p < 0.05 being statistically significant).

During the analysis, if certain data was not available for a particular patient, that patient was excluded from that specific group analysis. For example, if tumor grade was unavailable for a patient, that patient was not included when analyzing the relationship between tumor grade and APRI score. However, that patient was still included in the analysis of the remaining variables. Also, outliers were excluded from the analysis in of the laboratory parameters alkaline phosphatase, bilirubin, CA19-9, and INR with 5 times above the median being considered an outlier for alkaline phosphatase, CA19-9, and INR and 30 times above the median being considered an outlier for bilirubin. This led to exclusion of 5 values for alkaline phosphatase, 1 value for bilirubin, 16 values for CA19-9, and 1 value for INR.

## Results

### Demographic Data and APRI Score

In this study, the association between APRI score and age, sex, race, tumor type, and fibrosis etiology were determined. When looking at age, APRI score decreased at a rate of 0.01 with each additional year in age (r = -0.19, p = 0.02) (**Figure 1A**). However, given the small rate of change, this relationship between APRI score and age is not clinically significant.

**Figure 1:**
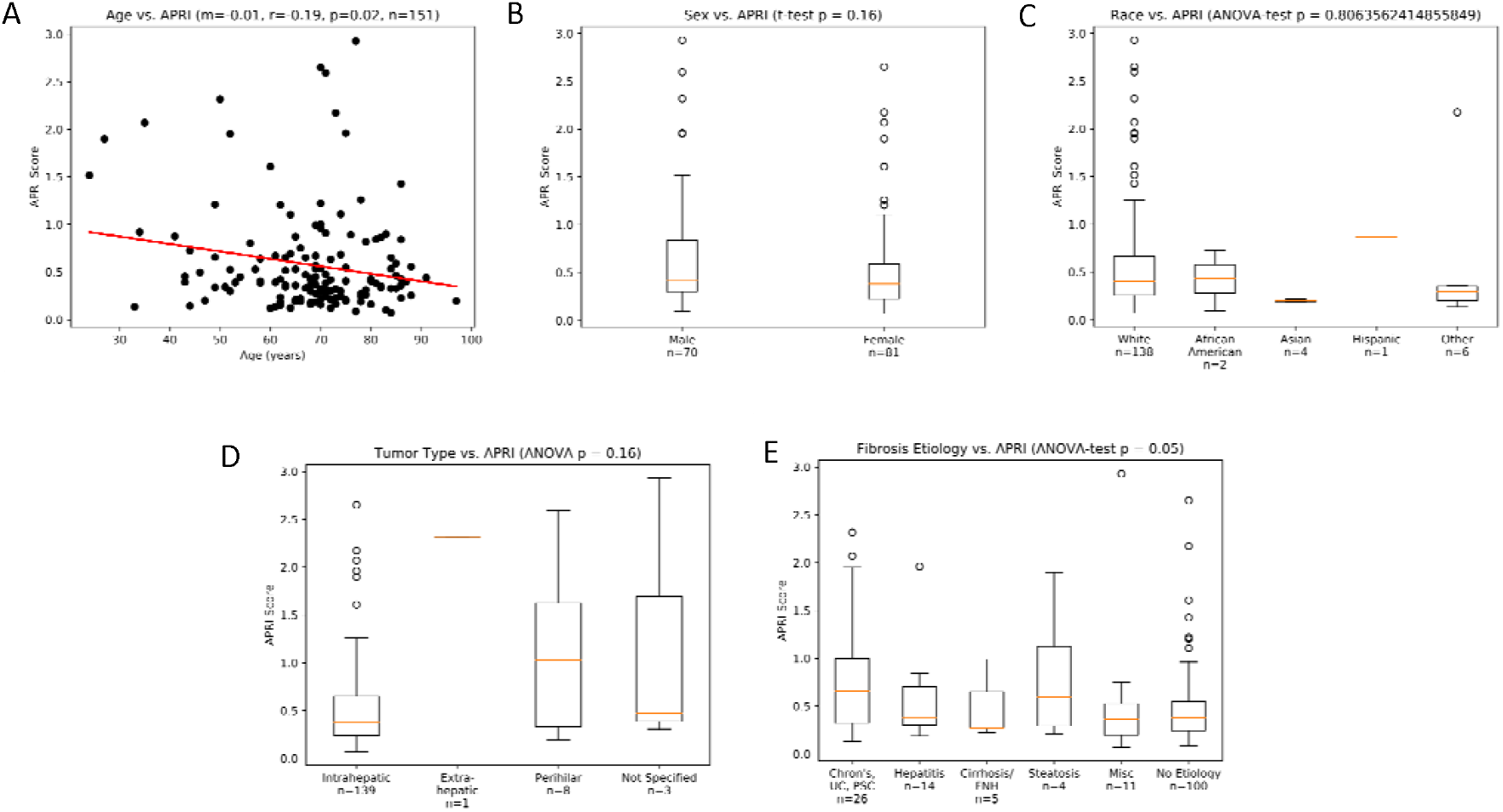
Clinical Characteristics and correlation with APRI score. (A) Scatter plot with line of best fit illustrating relationship between Age and APRI Score. (B) Boxplot illustrating variations in APRI score between sex (males vs. females). (C) Boxplot illustrating variations in APRI score in relation to Race. (D) Boxplot illustrating variations in APRI score among various tumor types. (E) Boxplot illustrating variation in APRI score among various fibrosis etiologies.

When looking at sex, there was no relationship between sex and APRI score. The median APRI score for males was 0.41 and for females was 0.38 with a 2-sample t test p-value of 0.16 (**Figure 1B**) (**Table 1**). There was also no relationship between race and APRI score. The median APRI score for white patients was 0.40, for African American patients was 0.43, for Asian patients was 0.20, for Hispanic patients was 0.90, and patients in the other category was - 0.29 with an ANOVA-test p-value of 0.81 (**Figure 1C**) (**Table 1**). It should be noted however that most of the patients in our study were white, and as such broader conclusions regarding impact of race on APRI score are not feasible from this dataset.

**Table 1:**
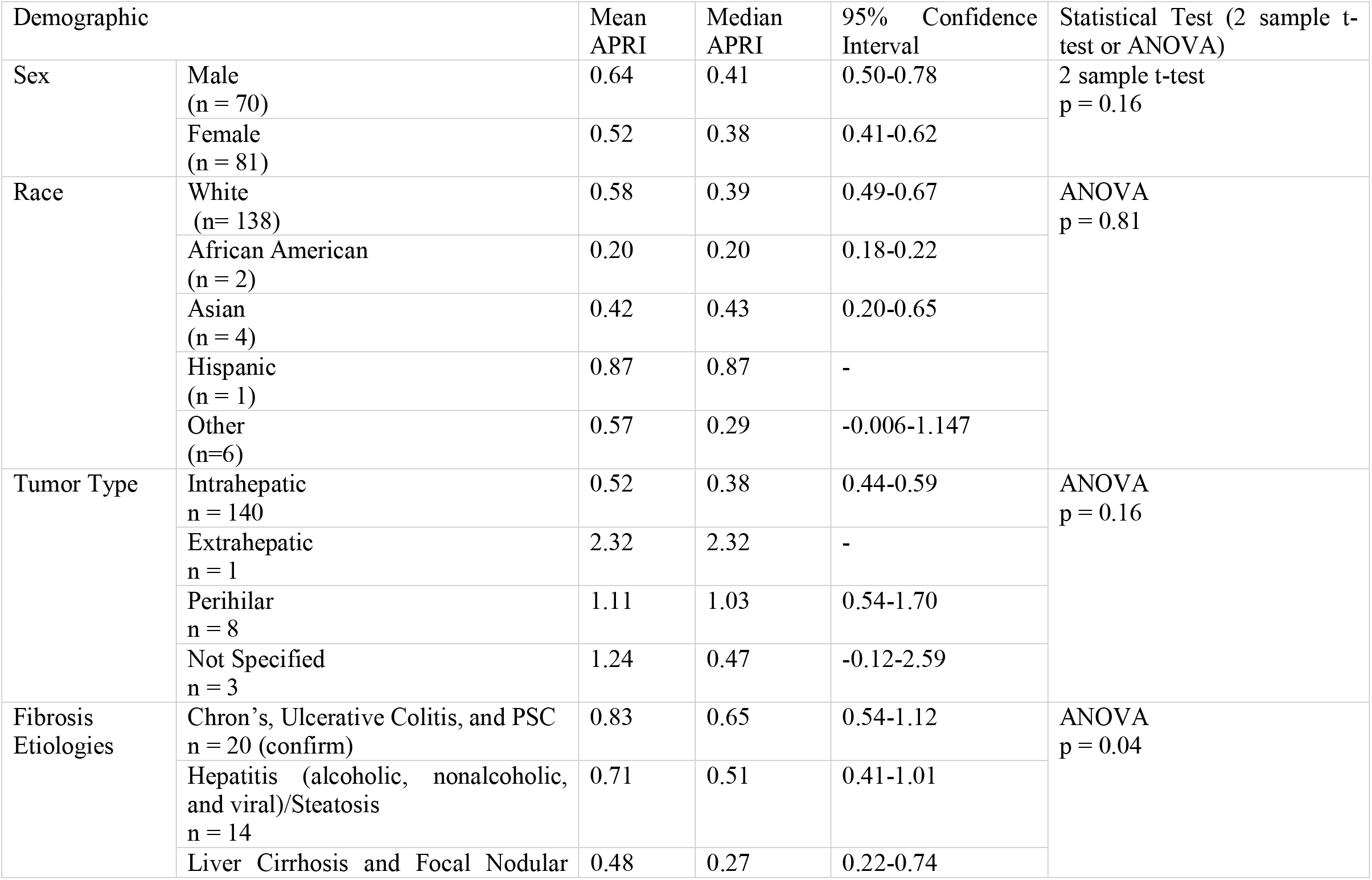
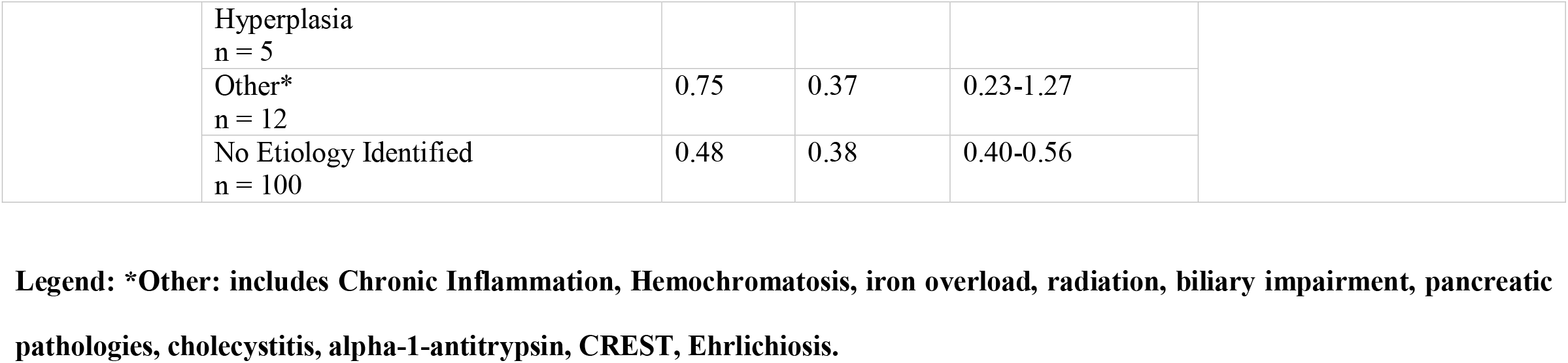
Sex, Race, Tumor Type, Fibrosis Etiology, and relationship to APRI score.

When looking at tumor type, 140 out of the 152 patients analyzed had intrahepatic cholangiocarcinoma and had a median APRI score of 0.38 (**Figure 1D**) (**Table1**). The remaining tumor types had no more than 3 patients in each category and thus meaningful conclusions could not be made regarding how tumor type affects APRI score. Regarding fibrosis etiology, the 20 patients with inflammatory bowel disease (ulcerative colitis or Crohn’s disease) or primary sclerosing cholangitis had a higher APRI score, with a median APRI score of 0.65, when compared to patients with other or no underlying etiologies. When looking at the remaining etiologies, the 14 patients with hepatitis (alcoholic, nonalcoholic, or viral) or steatosis had a median APRI score of 0.51, the 5 patients with liver cirrhosis or focal nodular hyperplasia had a median APRI score of 0.27, and the 12 patients with other causes of fibrosis had a median APRI score of 0.37 (**Figure 1E**) (**Table 1**). Most patients analyzed (100 out of 152) had no underlying fibrosis etiology and these patients had a median APRI score of 0.38.

### Laboratory Data and APRI Score

The relationship between APRI score and laboratory parameters including albumin, bilirubin, ALT, alkaline phosphatase, CA19-9, and INR were also determined.

After excluding outliers, when assessing the 144 patients with a recorded albumin level, the APRI score decreased at a rate of 0.35 for every 1 unit increase in Albumin (r = -0.35, p < 0.01) (**Figure 2A**). Among the 145 patients with a recorded bilirubin level, APRI score went up by 0.13 for every 1 unit increase in bilirubin (r = 0.23, p = 0.01) (**Figure 2B**). For the 129 patients with a recorded ALT, we found that APRI score went up by 0.1 for every 10 unit increase in ALT (r = 0.47, p < 0.01) (**Figure 2C**). Among the 145 patients with a recorded alkaline phosphatase level, there was a 0.18 increase in APRI score for every 100 unit increase in alkaline phosphatase (r = 0.41, p < 0.01).

**Figure 2:**
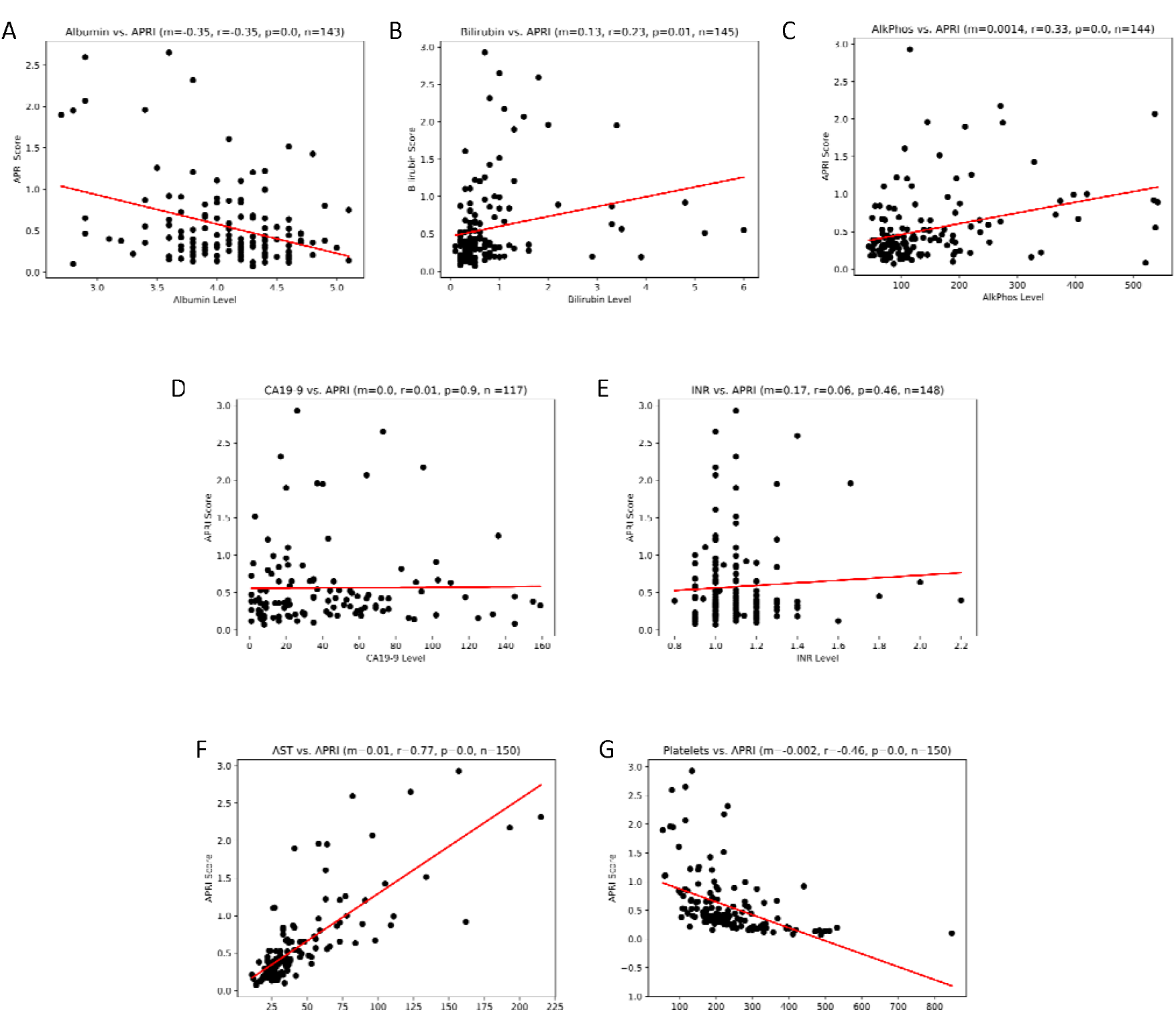
Laboratory Values and correlation with APRI Score. (A) Albumin. (B) Bilirubin. (C) Alkaline Phosphatase. (D) CA19-9 (E) INR. (F) AST. (G) Platelet count.

A total of 134 patients had a recorded CA19-9 level from which 16 outliers were excluded. From the remaining patients, CA19-9 did not correlate with APRI score in patients (m < 0.01, r = 0.01, p = 0.90). Similarly, there was no significant relationship between APRI score and INR among the 148 patients with a recorded INR (m = 0.17, r = 0.06, p = 0.46) (**Figure 2D and 2E**).

We also analyzed the relationship between APRI score with AST and platelet count. As would be expected, there was a strong correlation between AST and APRI (m = 0.013, r = 0.77, p < 0.01) and platelet count and APRI (m = - 0.002, r = -0.46, p < 0.01) (**Figure 2F and 2G**). These findings provide internal confirmation for validity of the APRI score given that platelet count and AST are components of APRI score determination.

### Tumor Pathology and APRI Score

The relationship between APRI score and tumor grade, N stage, T stage, presence of vascular invasion, presence of perineural invasion, tumor size, margin status, and margin width were also assessed.

When looking at tumor grade, the median APRI score for patients with grade G1 or G2 was 0.36 and for those G3 or G4 was 0.40 with a 2-sample t-test p = 0.86, meaning there was no relationship between APRI score and grade (**Figure 3A**). Regarding T stage, T3 and T4 had a slightly higher median APRI score of 0.52 when compared to the median APRI score for T1 and T2, which was 0.38 (2 sample t-test p = 0.01) (**Table 2**) (**Figure 3B**). However, only 19 patients in the cohort had either T3 or T4 stage compared to the 122 patients with either a T1 or T2 stage. Thus, a larger sample size for T3 and T4 stage would be needed to make more definitive conclusions about the relationship between T stage and APRI score. For N stage, the patient cohort consisted of tumors with either NX, N0 or N1. The median APRI score for NX was 0.36, for N0 was 0.38, and for N1 was 0.40 (one-way ANOVA p = 0.94) (**Figure 3C**).

**Table 2:** Tumor pathology and relationship to APRI score.

| Tumor Pathology Metric |  | Mean APRI | Median APRI | 95% Confidence Interval | Statistical Test (2 sample t-test or ANOVA) |
| --- | --- | --- | --- | --- | --- |
| Grade | G1+G2<br>(n = 94) | 0.51 | 0.36 | 0.41-0.60 | 2 sample t-test<br>p = 0.86 |
|  | G3+G4<br>(n = 47) | 0.59 | 0.40 | 0.43-0.75 |  |
| T Stage | T1+T2<br>(n = 122) | 0.51 | 0.38 | 0.43-0.59 | 2 sample t-test<br>p = 0.01 |
|  | T3+T4<br>(n = 19) | 0.82 | 0.52 | 0.49-1.15 |  |
| N Stage | NX<br>(n = 27) | 0.54 | 0.36 | 0.32-0.75 | ANOVA<br>p = 0.94 |
|  | N0<br>(n = 78) | 0.56 | 0.38 | 0.44-0.68 |  |
|  | N1<br>(n=37) | 0.53 | 0.40 | 0.40-0.66 |  |
| Vascular Invasion | Yes<br>(n = 40) | 0.58 | 0.44 | 0.44-0.71 | 2 sample t-test<br>p = 0.59 |
|  | No<br>(n = 106) | 0.53 | 0.36 | 0.43-0.63 |  |
| Perineural Invasion | Yes<br>(n = 30) | 0.68 | 0.47 | 0.47-0.88 | 2 sample t-test<br>p = 0.14 |
|  | No<br>(n = 89) | 0.50 | 0.36 | 0.40-0.60 |  |
| Margin Status | R0<br>n = 126 | 0.52 | 0.37 | 0.43-0.60 | R0 vs R1<br>2 sample t-test<br>p = 0.17 |
|  | R1<br>n = 18 | 0.70 | 0.53 | 0.42-0.98 |  |
|  | Not Specified n = 8 | 1.15 | 1.08 | 0.68-1.63 |  |

**Table 3:** Post-surgical outcomes and relationship to APRI score.

| Post-Resection Status |  | Mean APRI | Median APRI | 95% Confidence Interval | 2 sample t-test |
| --- | --- | --- | --- | --- | --- |
| 5-year Recurrence | Yes (n = 73) | 0.52 | 0.39 | 0.43-0.61 | p = 0.22 |
|  | No (n = 68) | 0.63 | 0.38 | 0.48-0.79 |  |
| 5-year Mortality | Yes (n = 54) | 0.54 | 0.39 | 0.43-0.64 | p = 0.39 |
|  | No (n = 92) | 0.61 | 0.39 | 0.48-0.73 |  |

**Figure 3:**
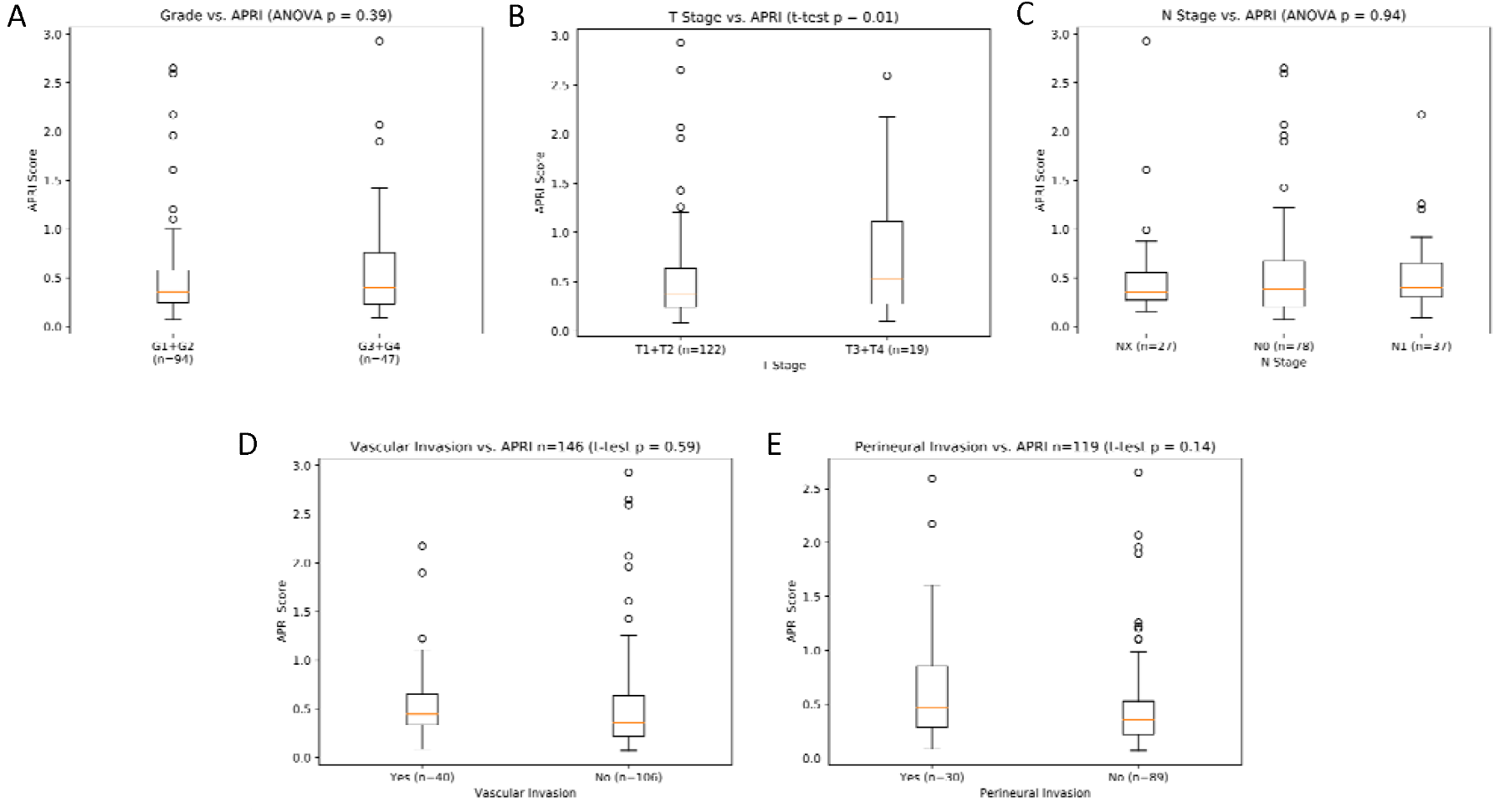
Tumor Grade, Stage, Vascular Invasion, and Perineural Invasion related to APRI Score. (A) Grade. (B) T Stage. (C) N Stage. (D) Vascular Invasion. (E) Perineural Invasion.

APRI score also did not have a relationship with vascular invasion and perineural invasion. The median APRI score for patients with vascular invasion was 0.44 and for those without vascular invasion was 0.36 (2-sample t-test p = 0.59) (**Figure 3D**). Similarly, the median APRI score for patients with perineural invasion was 0.47 and for those without it was 0.36 (2-sample t-test p = 0.14) (**Figure 3E**). There was also no relationship between margin status and APRI score. 126 out of the 152 patients with a margin status of R0 had a median APRI score of 0.37 and the 18 patients with a margin status of R1 had an APRI score of 0.53 (2 sample t-test p = 0.17). The remaining 8 patients did not have a reported margin status.

Pathological tumor size and margin width were also not related to APRI score. Pathology size with a m = -0.01, r = 0.12, and p = 0.61 when correlated with APRI score (**Figure 4A**). Similarly, margin width with a m = 0.01, r = 0.12, p = 0.16 when correlated with APRI score (**Figure 4B**).

**Figure 4:**
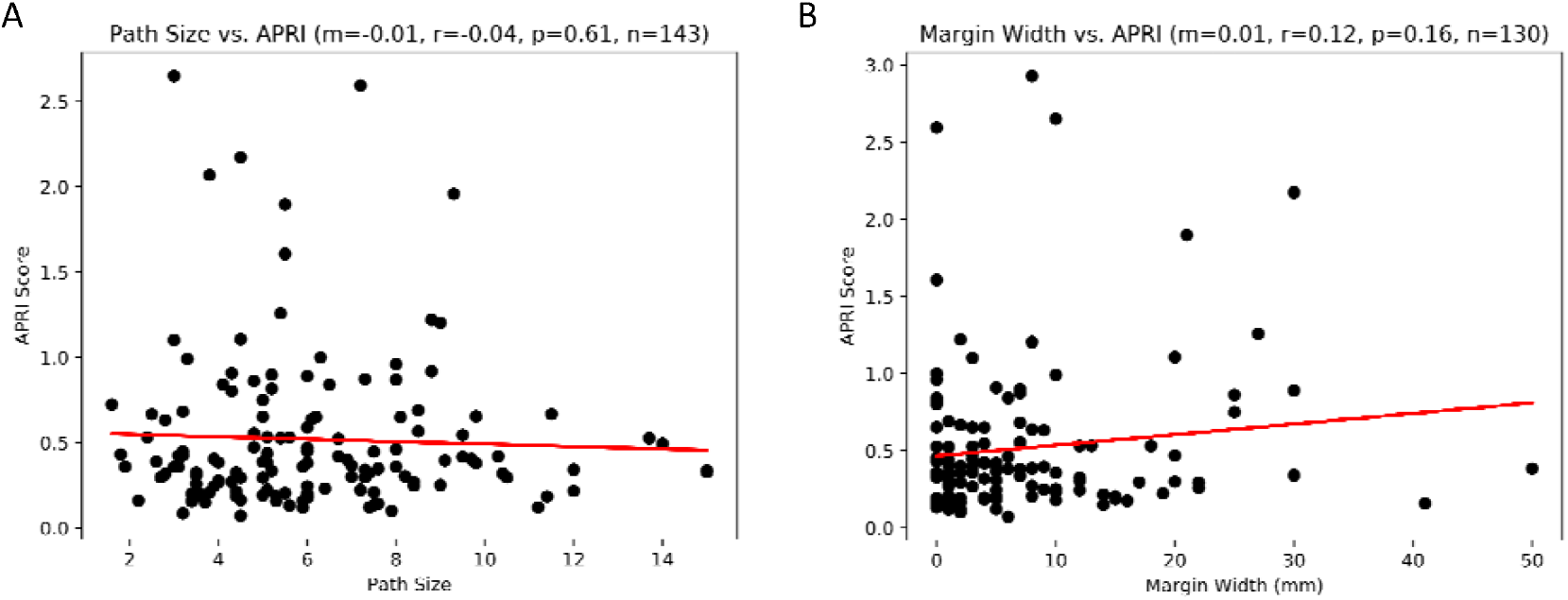
Pathology Size and Margin Width related to APRI Score. (A) Pathology Size. (B) Margin Width.

### Post-Surgical Outcomes and APRI Score

Post-surgical outcomes that were analyzed included post-surgical recurrence and post-surgical death. The corresponding APRI scores were subsequently used to assess whether there is a relationship with these outcomes.

When looking at tumor recurrence, the median APRI score for patients with recurrent disease within 5 years was 0.39 and the median APRI score for those without recurrence was (2-sample t-test p = 0.22) (**Figure 5A**). Among patients with recurrent disease, there was no correlation between days to recurrence and APRI score (m < 0.01, r = 0.05, p = 0.67) (**Figure 5B**).

**Figure 5:**
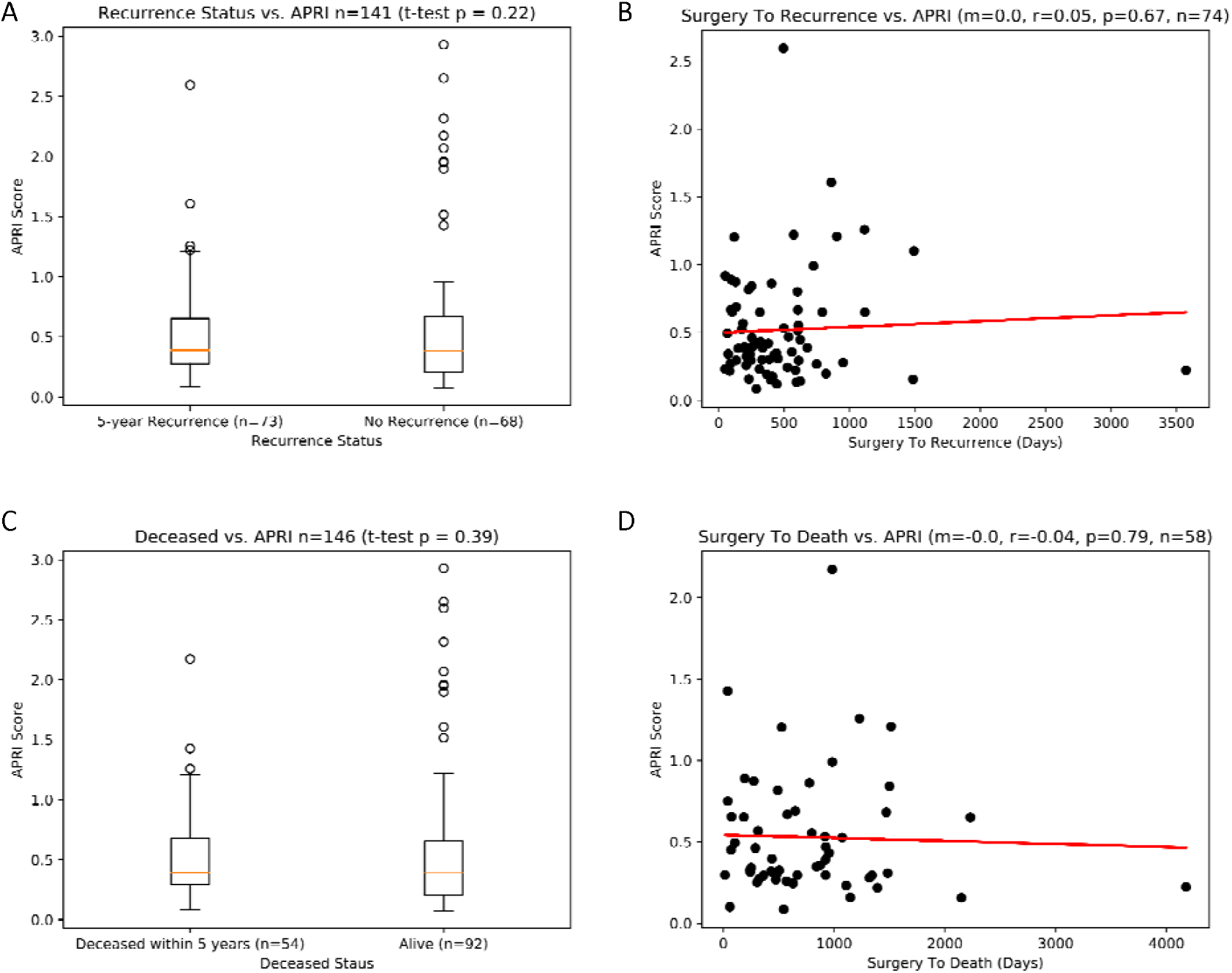
Post-resection Tumor Recurrence, Death, and APRI Score. (A) Recurrence Status. (B) Days to Recurrence. (C) Deceased Status. (D) Days to Death

When looking at deceased status, the median APRI score for patients deceased within 5 years of surgery was 0.39 and the median APRI score for patients who were not deceased was also 0.39 (2-sample t-test p = 0.39) (**Figure 5C**). Among patients who were deceased, there was no correlation between days to death and APRI score (m < -0.01, r = -0.04, p = 0.80) (**Figure 5D**).

## Discussion

Given that one of the most common drivers of cholangiocarcinoma pathogenesis is chronic inflammation of the liver and biliary tree, we hypothesized that a marker of liver fibrosis, such as the APRI score, could have been a useful marker for prognosticating post-surgical outcomes.^3^ However, the results of our study illustrate this to not be the case.

In our analysis, we found no relationship between an array of demographic factors including age, sex, race, tumor type and APRI score. However, this may be due to the lack of diversity in our patient cohort. This is especially true for races as most of the patients in our study were white. However, when looking at underlying fibrosis etiologies, patients with IBD or PSC had a higher APRI score when compared to other etiologies. This is likely because IBD and PSC are inflammatory processes leading to increased liver fibrosis and thus a higher APRI score. However, it should be noted that only 26 out of the 152 patient we analyzed had an IBD or PSC and so no meaningful conclusions can be made due to the limited sample size. We did find a relationship between laboratory parameters including ALT, bilirubin, and alkaline phosphatase and APRI score. However, although these findings may suggest a relationship between APRI score and liver function/inflammation, it does not make the case for its use as a prognostic tool in cholangiocarcinoma patients.

We ascertained that APRI score did not correlate with tumor pathology and post-surgical outcomes. We found no relationship between APRI score and tumor grade, stage, presence of vascular invasion, and presence of perineural invasion—all characteristics that have shown to be related to patient outcomes.^2, 5, 7, 8^ Moreover, we also found that APRI score was not related to post-surgical disease recurrence nor mortality. There can be several reasons for this discrepancy. Although chronic inflammation is thought to be a potential underlying driver of the development of cholangiocarcinoma, it may not be the sole driver of disease.^3, 16^ Other factors, like genetic, epigenetic, and aberrant signaling pathways may play a larger role disease pathogenesis compared to chronic inflammation.^16^ Thus, if chronic inflammation is not the primary driver of disease, then measures like APRI score would not be effective in prognosticating disease, as we have noted in our study.

However, this understanding of the pathogenesis of cholangiocarcinoma does not rule out the utility of APRI score in other settings as has been shown in prior studies. Rather, APRI score may still be useful in patients with underlying inflammatory etiologies of cholangiocarcinoma, like PSC, hepatitis virus, and liver cirrhosis, especially given that we found IBD and PSC patients to have higher APRI scores when compared to other groups in our cohort. Moreover, this has also been found to be the case in studies looking at the utility of APRI score in prognosticating hepatocellular carcinoma.^17^ A study conducted by Mai et al showed that a combination of aminotransferase-platelet ratio index (APRI) and albumin-bilirubin (ALBI) scores correlated with post-surgical liver failure in patients with hepatitis B virus related hepatocellular carcinoma.^17^ This would make sense given that APRI score has shown to be a marker for liver fibrosis and since HBV mediated HCC is largely driven by inflammation, more severe fibrosis should relate to worse outcomes.

Epidemiologic factors also play a role when assessing the utility of APRI score. In Asian countries, like China and South Korea, inflammatory etiologies of cholangiocarcinoma are more prevalent due to the higher incidence of liver flukes, parasites, and HBV/HCV.^16^ On the other hand, in Western countries, the prevalence of these inflammatory etiologies is much lower.^16^ Since our study was conducted among patients in the United States, APRI score did not prove useful as a prognostic tool as most patients did not have underlying inflammatory etiologies. However, if the utility of APRI score was assessed among patients in high-risk regions where inflammatory etiologies are more common, then APRI score may have proven useful.

Our study did have some limitations. This study was a retrospective analysis and therefore is associated with weaknesses related to such studies. This study was a single center study and as such population bias and referral bias would be factors that would be associated with this effort. Moreover, although we were able to discern underlying etiologies of disease in some patients, most patients in our cohort had no identifiable underlying etiology of their cholangiocarcinoma as the etiology of many cases are multifactorial and not readily discernable. Thus, we were unable to make meaningful conclusions regarding the relationship of APRI score on outcomes among the different subgroups of etiologies for cholangiocarcinoma in this study. Future studies could look at the utility of APRI score in prognosticating disease in patients within these subgroups.

## Conclusion

In conclusion, we found that APRI score was not useful to prognosticate post-surgical outcomes in cholangiocarcinoma. These findings were contrary to our initial hypothesis that chronic inflammation could be a key driver of outcomes in cholangiocarcinoma pathogenesis.^3^ However, these findings suggest that the pathogenesis of cholangiocarcinoma cannot be explained by chronic inflammation alone. Rather, a combination of factors like genetic, epigenetic, and epidemiological factors may all influence the pathogenesis of disease.^16^

## Data Availability

All data produced in the present study are available upon reasonable request to the authors

